# Protein Classifier for Thyroid Nodules Learned from Rapidly Acquired Proteotypes

**DOI:** 10.1101/2020.04.09.20059741

**Authors:** Yaoting Sun, Sathiyamoorthy Selvarajan, Zelin Zang, Wei Liu, Yi Zhu, Hao Zhang, Hao Chen, Xue Cai, Huanhuan Gao, Zhicheng Wu, Lirong Chen, Xiaodong Teng, Yongfu Zhao, Sangeeta Mantoo, Tony Kiat-Hon Lim, Bhuvaneswari Hariraman, Serene Yeow, Syed Muhammad Fahmy bin Syed Abdillah, Sze Sing Lee, Guan Ruan, Qiushi Zhang, Tiansheng Zhu, Weibin Wang, Guangzhi Wang, Junhong Xiao, Yi He, Zhihong Wang, Wei Sun, Yuan Qin, Qi Xiao, Xu Zheng, Linyan Wang, Xi Zheng, Kailun Xu, Yingkuan Shao, Kexin Liu, Shu Zheng, Ruedi Aebersold, Stan Z. Li, Oi Lian Kon, N. Gopalakrishna Iyer, Tiannan Guo

## Abstract

Up to 30% of thyroid nodules cannot be accurately classified as benign or malignant by cytopathology. Diagnostic accuracy can be improved by nucleic acid-based testing, yet a sizeable number of diagnostic thyroidectomies remains unavoidable. In order to develop a protein classifier for thyroid nodules, we analyzed the quantitative proteomes of 1,725 retrospective thyroid tissue samples from 578 patients using pressure-cycling technology and data-independent acquisition mass spectrometry. With artificial neural networks, a classifier of 14 proteins achieved over 93% accuracy in classifying malignant thyroid nodules. This classifier was validated in retrospective samples of 271 patients (91% accuracy), and prospective samples of 62 patients (88% accuracy) from four independent centers. These rapidly acquired proteotypes and artificial neural networks supported the establishment of an effective protein classifier for classifying thyroid nodules.

## INTRODUCTION

Advances in imaging technology and liberal screening practices have identified thyroid nodules in up to 50% of the general population, but only a small minority (7-15%) eventually prove to be malignant by histology, and even fewer among these are clinically relevant (Burman and Wartofsky, 2015; Jameson, 2012). Beyond clinical assessment and ultrasonography, fine needle aspiration (FNA) followed by cytopathology is considered the most reliable pre-surgical technique for differentiating benign from malignant thyroid tumors (Burman and Wartofsky, 2015; Faquin et al., 2011). Yet up to one-third of thyroid nodules are deemed indeterminate by FNA-cytopathology (Alexander et al., 2012), and surgery remains the only option for accurate diagnosis. However, the majority of thyroid surgeries are diagnostic procedures undertaken to exclude thyroid cancer, of which no more than 25% accomplish any therapeutic purpose (Ahn et al., 2014). Patients whose thyroid glands are removed in part or entirely often require daily and lifelong thyroxine-replacement therapy and medical monitoring. Given that only 10% of resected glands prove to be malignant, the current clinical approach results in substantial over-treatment with unwarranted surgical risks for patients who could otherwise be treated conservatively (Vaccarella et al., 2016).

Molecular tests adjunctive to FNA-cytopathology have focused on RNA expression or DNA mutational profiling of aspirates obtained prior to surgery, using small quantities of RNA or DNA that can be amplified (Eszlinger et al., 2017). The Afirma® gene expression classifier (GEC) determines RNA abundances of 142 genes with high sensitivity and negative predictive value (NPV) of up to 92% and 93%, respectively. However, positive predictive value (PPV) and specificity for diagnosing malignancy are only 47% and 52%, respectively (Alexander et al., 2012). The latest version of Afirma® called GSC has achieved a sensitivity of 91%, but the specificity was only 68% (Patel et al., 2018). The ThyroSeq (v3.0) test combines mutation and expression profiling for better accuracy (Nikiforova et al., 2013), but has other limitations i.e. the need for fresh tissue samples with un-degraded RNA, and test data processing by central ‘black box’ analysis. A recent review of the GEC test showed sensitivities of 83-100% but low overall specificity, ranging from 10 to 52% across multiple centers. This may reflect variability in sample composition, issues with tissue quality and the fragility of nuclei acid-based testing in general (Wang and Sosa, 2018). While nucleic acid-based approaches continue to be refined, for example with successive iterations of ThyroSeq panels, there is evident scope for alternative approaches to address this diagnostic dilemma.

Until recently, proteomics-based analyses were limited to large tissue quantities and fresh/snap frozen samples. Proteotyping hundreds of biopsy-level tissue samples from clinical cohorts remains unfeasible with conventional methods. We developed a pressure cycling technology (PCT) protocol for proteomic analysis of tissue biopsy samples (Guo et al., 2015) which can be performed on minimal amounts (0.2–1 mg) of fresh-frozen tissue samples (Shao et al., 2016; Shao et al., 2015). The method was recently extended to generate high quality proteome data from formalin-fixed, paraffin-embedded (FFPE) tissue samples (Zhu et al., 2019), and in this study on fine-needle thyroid gland aspirates. Coupled with Sequential Window Acquisition of all Theoretical fragment ions (SWATH) mass spectrometry (MS), a data-independent acquisition (DIA)-MS method (Gillet et al., 2012), this technique now permits practical proteomic analysis of biopsy size FFPE tissue samples at high sample throughput. In this study we apply this technology to analyze a large number of tissue samples and show that the high quality proteotype data generated a robust panel of protein markers which differentiates benign from malignant thyroid disease with high ‘rule in’ and ‘rule out’ accuracy.

## RESULTS

### Study design and clinical characteristics

We applied PCT-DIA on a total of 931 nodules from 911 patients using tissue cores (1 mm diameter with 0.5-1 mm thickness) punched out from regions of interest marked on FFPE tissue blocks, or from fine-needle aspiration (FNA) biopsies. The samples comprise (i) a discovery set from Singapore General Hospital (n = 579 nodules) where histopathological diagnoses were confirmed on central review by a board-certified pathologist; and (ii) independent test sets from four hospitals in China consisting of retrospective test sets of FFPE samples (n = 288 nodules) and prospective test set of FNA biopsies (n = 64 nodules) (Figure 1A). The discovery set comprised FFPE samples from 40 normal thyroid tissues (N), 203 multinodular goiters (MNG), 137 follicular adenomas (FA), 75 follicular carcinomas (FTC) and 124 papillary carcinomas (PTC) (Table 1; Table S1). For subsequent analyses, these were divided into benign (comprising N, MNG and FA) and malignant (comprising FTC and PTC) thyroid nodules. For each nodule in the discovery set, three punches were obtained from the region of interest as biological replicates. In total, we analyzed 1,725 samples randomly distributed into 121 batches to minimize batch effects (Figure 1B) using 45 min DIA-MS, and an additional 56 randomly selected samples as technical replicates. These technical replicates distributed randomly in the first 84 batches for the training sample set. Although higher proteomic depth could be obtained with a longer LC gradient, we adopted a reasonably fast analysis time to minimize batch effects without substantial compromise of proteome depth (Sun, 2020), thus facilitating effective downstream machine learning to establish a robust classifier.

**Table 1.** Clinico-pathologic characteristics of the study cohorts.

|  | Discovery dataset | Independent test datasets |  | All |
| --- | --- | --- | --- | --- |
|  |  | Retrospective test datasets | Prospective test dataset |  |
| **Total no.** |  |  |  |  |
| Clinical centers | 1 | 3 | 1 | 5 |
| patients | 578 | 271 | 62 | 911 |
| nodules | 579 | 288 | 64 | 931 |
| FFPE punches | 1725 | 288 | 0 | 2013 |
| Fine needle aspiration biopsies | 0 | 0 | 64 | 64 |
| DIA files | 1781 | 576 | 64 | 2421 |
| **Histopathology diagnosis** |  |  |  |  |
| Normal thyroid tissues | 40 (6.9%) | 16 (5.6%) | 0 (0.0%) | 56 (6.0%) |
| Multinodular goiter | 203 (35.1%) | 44 (15.3%) | 13 (20.3%) | 260 (27.9%) |
| Follicular adenoma<sup>b</sup> | 137 (23.7%) | 84 (29.2%) | 5 (7.8%) | 226 (24.3%) |
| Follicular thyroid carcinoma<sup>b</sup> | 75 (13.0%) | 52 (18.1%) | 0 (0.0%) | 127 (13.6%) |
| Papillary thyroid carcinoma | 124 (21.4%) | 92 (31.9%) | 46 (71.9%) | 262 (28.1%) |
| **Diagnosis age** |  |  |  |  |
| Mean | 52.8 | 47.5 | 43.6 | 50.6 |
| Median | 54 | 49 | 44 | 52 |
| Range | 13-85 | 15-82 | 24-74 | 13-85 |
| < 18 y | 7 (1.2%) | 1 (0.4%) | 0 (0.0%) | 8 (0.9%) |
| 18-55 y | 288 (49.8%) | 173 (63.8%) | 48 (77.4%) | 509 (55.9%) |
| ≥55 y | 283 (49.0%) | 97 (35.8%) | 14 (22.6%) | 394 (43.2%) |
| **Gender** |  |  |  |  |
| Female | 433 (74.9%) | 191 (66.3%) | 54 (87.1%) | 678 (74.4%) |
| Male | 145 (25.1%) | 80 (27.7%) | 8 (12.9%) | 233 (25.6%) |
| **Nodule size<sup>a</sup> on ultrasonography** |  |  |  |  |
| Mean | 3.3 | 2.9 | 2.5 | 3.1 |
| Median | 3.0 | 2.6 | 2.2 | 2.9 |
| Range | 0.2-13.0 | 0.2-8.9 | 0.6-5.5 | 0.2-13.0 |
| < 1 cm | 15 (2.8%) | 37 (14.0%) | 2 (3.1%) | 54 (6.3%) |
| 1~4 cm | 342 (64.5%) | 143 (54.0%) | 53 (82.8%) | 538 (62.6%) |
| ≥4 cm | 173 (32.6%) | 85 (32.1%) | 9 (14.1%) | 267 (31.1%) |

<sup>a</sup>Nodule size information of nine patients in discovery dataset and seven patients in test dataset were not record.
<sup>b</sup>Hurthle cell adenoma and carcinoma were allocated into follicular thyroid adenoma and carcinoma, respectively.

**Figure 1.**
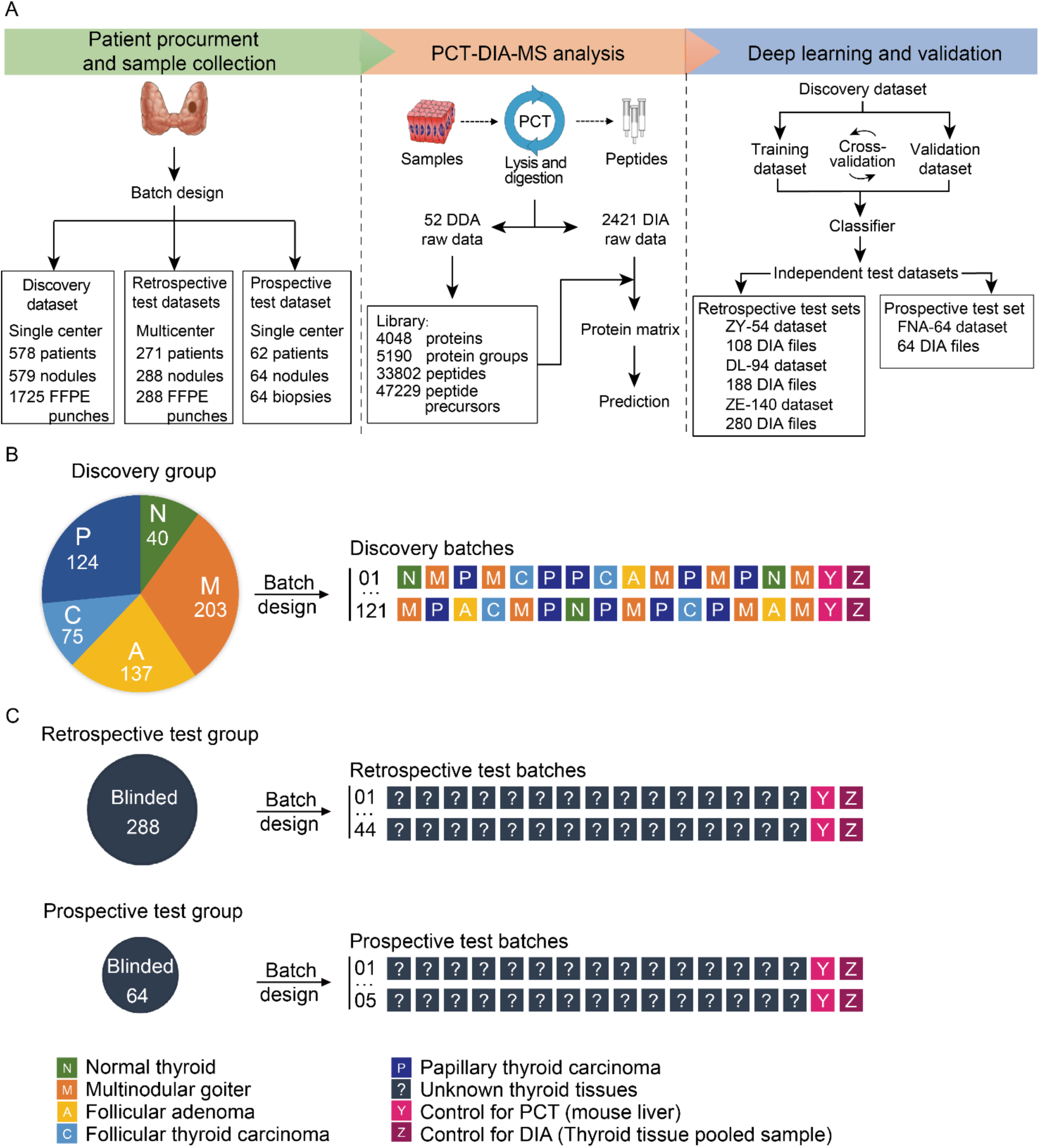
Schematic view of the study and batch design. (A) The project design and workflow of the FFPE PCT-DIA pipeline. (B-C) Batch design (B) The discovery group of 579 thyroid nodules from 578 patients consisted of 40 normal thyroid, 203 multinodular goiter, 137 follicular thyroid adenoma, 75 follicular thyroid carcinoma and 124 papillary thyroid carcinomas with unblinded diagnoses. Each nodule was represented by three cores as biological replicates. 1,781 thyroid FFPE punches and 56 technical replicates were randomly allocated into 121 discovery batches in order to minimize batch effect for this large-scale sample preparation. (C) The independent test group contained two parts, one is retrospective test group and the other is prospective test group. Retrospective test group included 288 thyroid nodules of blinded diagnoses from 271 patients. Each nodule was analyzed in technical duplicates, without biological replicates. A total of 288 FFPE cores and 288 corresponding technical duplicates were divided into 44 batches for analysis. Prospective test group contained 64 fine needle aspiration biopsies of thyroid nodules from 64 patients which were divided into 5 batches. Each batch consisted of 15 thyroid samples, one mouse liver sample and one pooled thyroid sample.

### Global proteomic profiling of thyroid nodules

To analyze the DIA data, we built a specific thyroid-tissue spectral library from FFPE tissues containing 52 DDA files (Table S2) using a 120-min liquid chromatography (LC) gradient on a Q Exactive Hybrid Quadrupole-Orbitrap mass spectrometer. Relatively long gradient was adopted for DDA analysis to maximize the isolation of peptide precursors. We constructed a spectral library containing 33,802 peptides from 5,190 protein groups using our previously established computational pipeline (Zhu, 2020). The number of proteins in this library is relatively smaller than that of other tissue types analyzed using a similar workflow (Guo et al., 2015; Shao et al., 2019; Zhu, 2020; Zhu et al., 2019) or other proteomic protocols (Jiang et al., 2019; Sinha et al., 2019; Zhang et al., 2016) as thyroid tissue contains a high abundance protein, thyroglobulin, which weakens the signal for the rest of the proteome (M et al., 2018). Using OpenSWATH (v2.0) and our thyroid library, we analyzed 1,781 DIA maps (1,725 FFPE cores and 56 technical replicates). We identified and quantified 30,915 peptides from 3,708 high-confidence proteotypic proteins (Table S3). Details on quality control and reproducibility (Figure S1) are detailed in the respective supplementary sections.

From these, we computed the average intensities of 2,617 proteotypic proteins which were quantified with high confidence for each thyroid nodule, as visualized in a tissue-type supervised heatmap (Figure 2A). Malignant tissue samples expressed higher number of proteins than benign samples, indicating more diverse proteome of tumor cells. To check whether the thus acquired proteotypes classifies different tissue types, we applied uniform manifold approximation and projection (UMAP) algorithm to visualize the 579 proteotypes from five tissue types (Figure 2B). The plot shows that PTC samples are well isolated from the rest, indicating PTC samples are vastly different in terms of proteome expression. We then grouped N, MNG and FA as benign group, while FTC and PTC as the malignant group (Figure 2C). The UMAP visualization shows that malignant samples are well resolved from benign samples with some overlap. We further narrowed our focus to the N and MNG groups and found that their proteotypes share high degree of similarity (Figure 2D). Not surprisingly, FA exhibited significant overlap with both benign and malignant subsets, particularly between FA and FTC (Figure 2E), corroborating biological overlap between the two pathologies, while there were sufficient features that could distinguish FTC from PTC (Figure 2F). FA and FTC could not be separated neither (Figure 2E). Pair-wise comparison of each two groups are shown in Figure S2. The above analyses show that the proteotype maps thus measured reasonably reflected the clinical phenotype of these samples.

**Figure 2.**
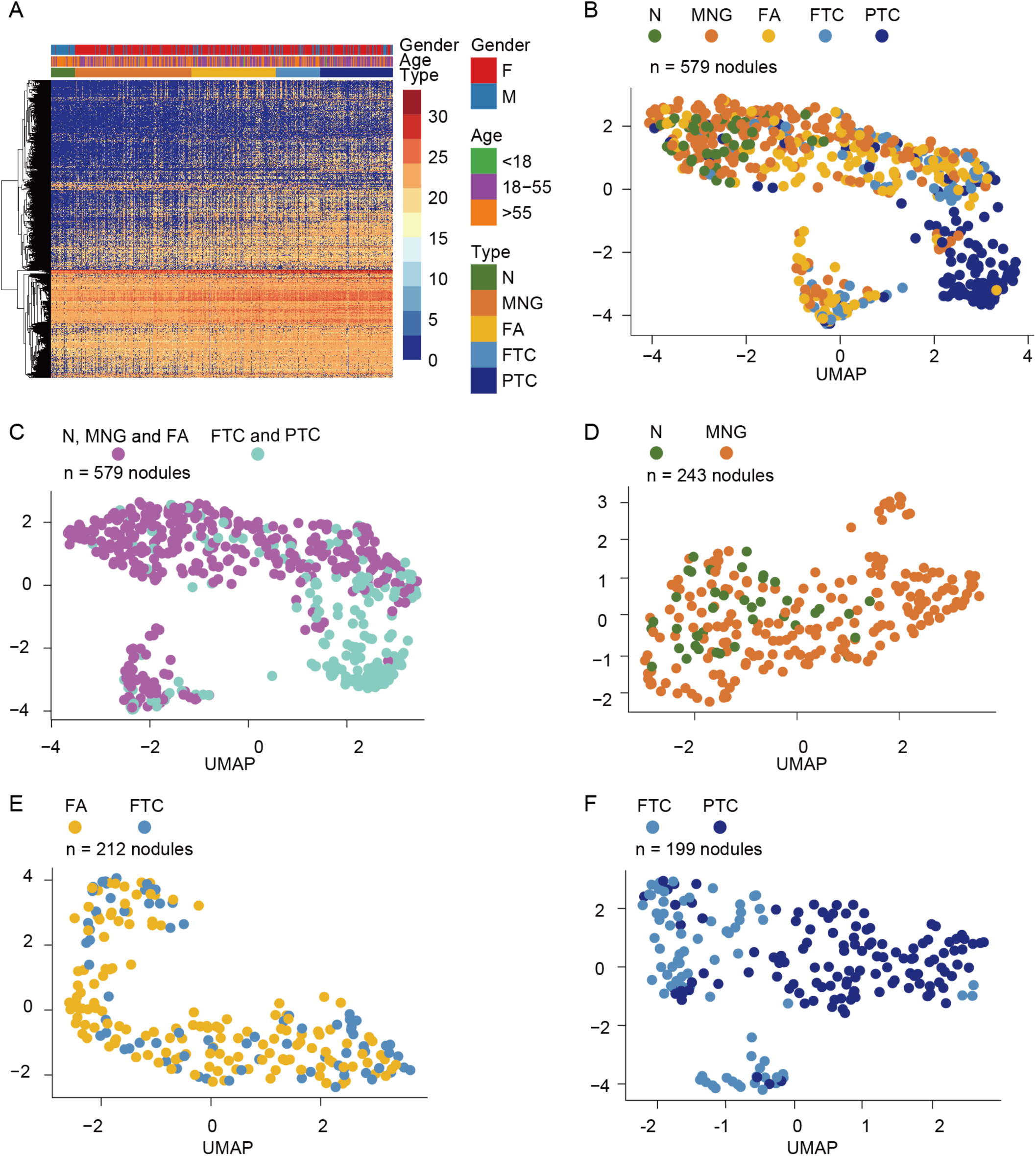
Global thyroid proteome profile. (A) Heatmap showing global protein expression profiles of the five main histological types of thyroid tissues. 579 thyroid tissues from 578 patients are in columns and 2,617 proteins (missing value <90%) in rows. The color bar indicates the intensity of the proteins. (B-F) UMAP plots showing global snapshots comparing the indicated types thyroid tissues using 2,617 proteins for (B) all subtypes; (C) benign versus malignant; (D) only benign; (E) FA versus FTC; and (F) only malignant tissue types.

### Feature selection and classifier development

To derive a protein-based signature to differentiate benign from malignant thyroid tumors, we developed a customized feature selection process combined with artificial neural network algorithms based on the discovery dataset of 500 samples (Figure 3A). The rest 79 samples serve as internal validation set. Here we limited the number of selected features to no more than 20 so that they may be practically measured by targeted proteomics or antibodies in clinic. We paid special attention to maximize the specificity while keeping the sensitivity above 90% since the major problem of thyroid nodule evaluation is over-diagnosis. Briefly, protein features selected by a genetic algorithm (Mitchell, 1998) combined with 5-fold cross validation led us to identify a classifier-panel of 14 proteins (Table 2) to separate benign and malignant nodules. Individual protein expression levels are shown in Figure 4. Ten of these proteins have been previously reported in thyroid cancers, including annexin A1 (Ciregia et al., 2016), galectin-3 (Bartolazzi et al., 2018), SH3 domain-binding glutamic acid-rich-like protein 3 (Martínez-Aguilar et al., 2016), alpha 2-HS glycoprotein (Farrokhi Yekta et al., 2018), myosin heavy chain 9 (Wang et al., 2017), phosphatidylethanolamine-binding protein 1 (Kim et al., 2010), clusterin (Kashat et al., 2010), LDL receptor related protein 2 (He et al., 2020), calreticulin (Schürch et al., 2019) and moesin (Smith et al., 2019). In addition, this list also includes two proteins involved thyroid functions, including tubulin folding cofactor A (Figliozzi et al., 2017) and histone cluster 1 H1 family member c (Brix et al., 1998). No previous association with thyroid disease has been reported for the remaining two (Thy-1 cell surface antigen and sialic acid acetylesterase).

**Table 2.**
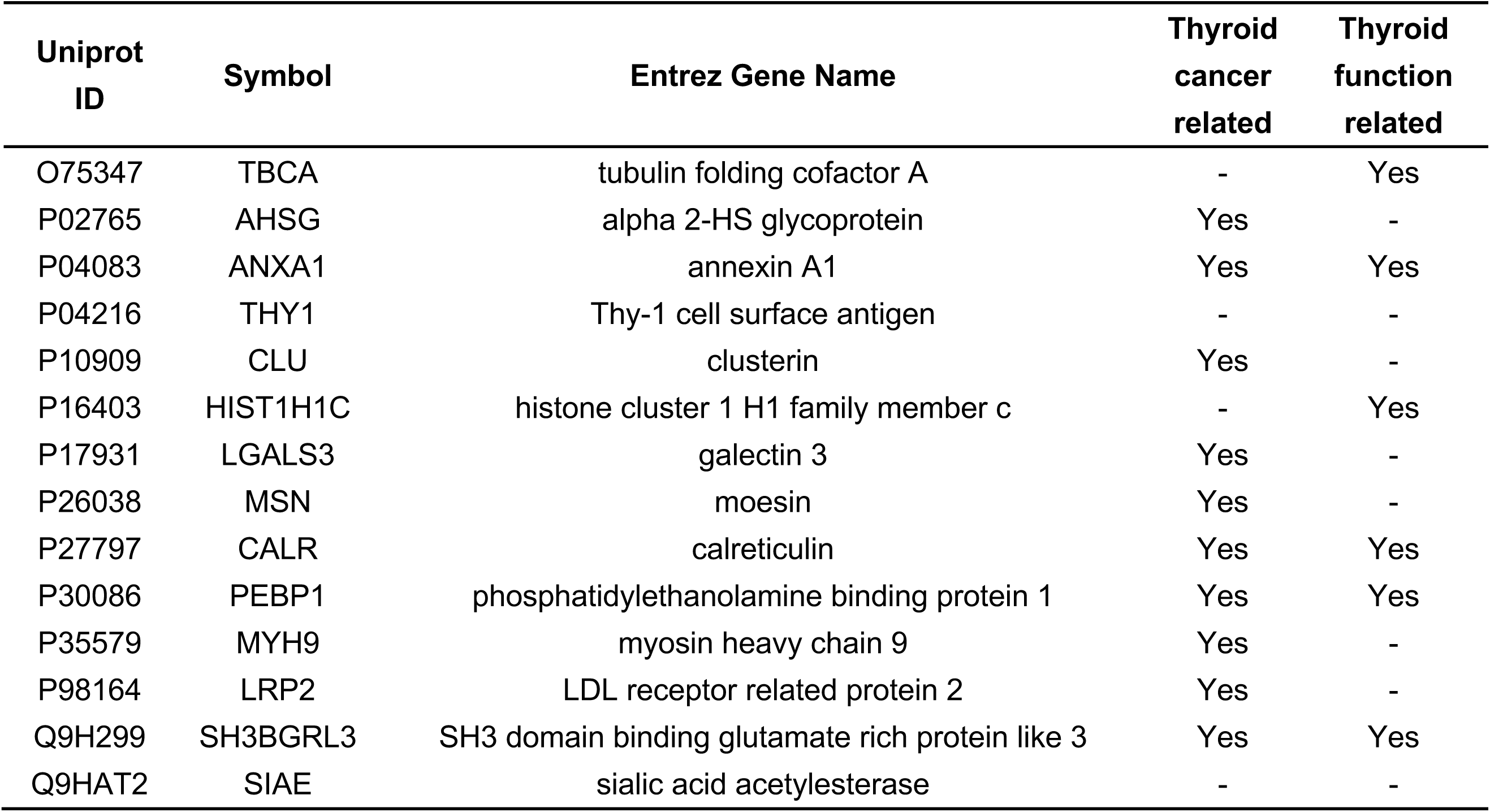
Fourteen proteins selected by genetic algorithm, functional pathways and previously known associated thyroid physiology or pathology.

**Figure 3.**
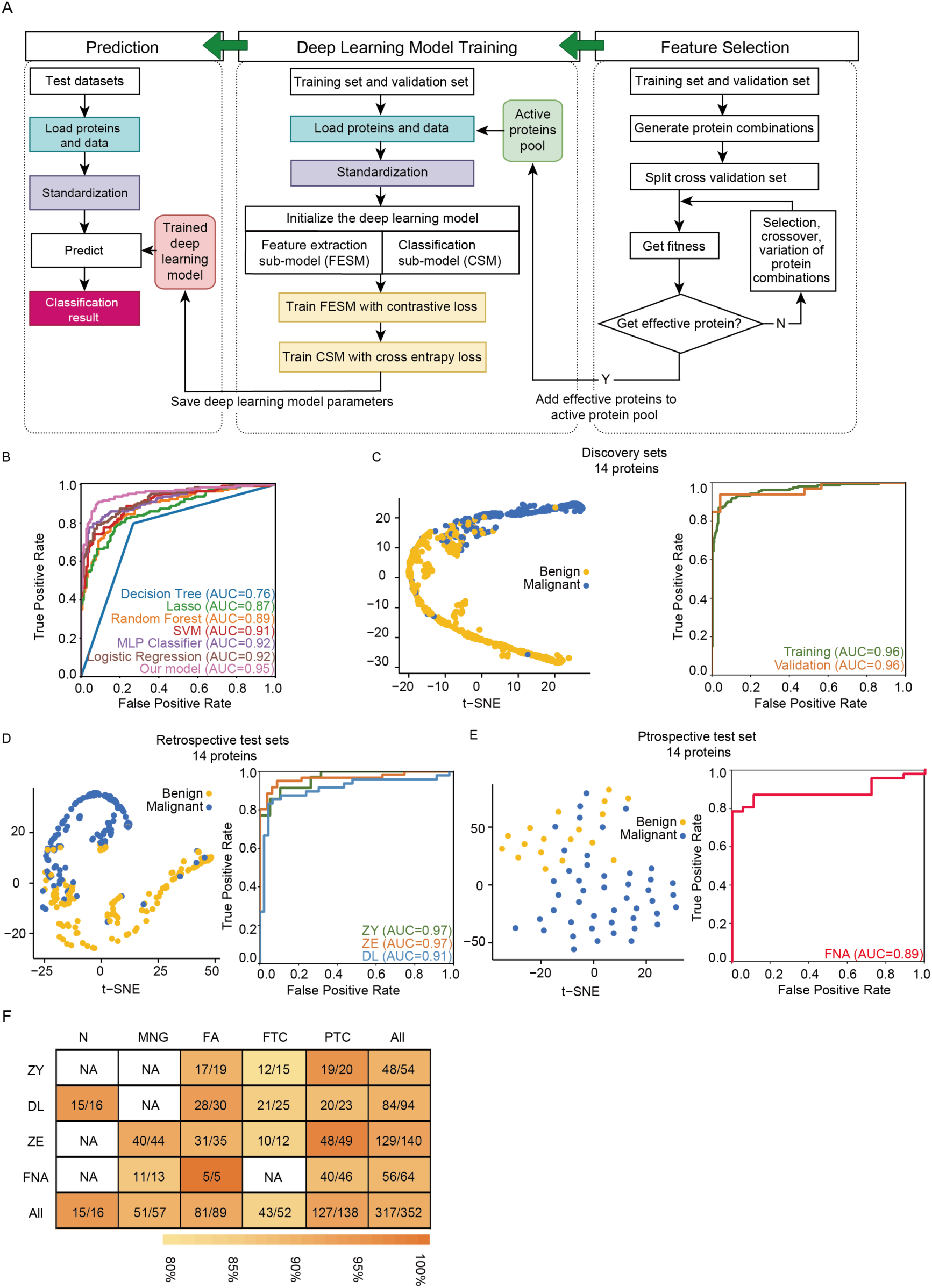
Classifier development, performance testing and validation in independent blinded datasets. (A) Schematic of principal classifier model (details in Supplementary Methods). (B) ROC plots of seven different machine learning model under 14 selected features. (C) t-SNE plot showing the separation between benign malignant groups in the discovery set using 14 protein features with latent space; and ROC plots of the training and validation sample subgroups of the discovery set. (D-E) t-SNE and ROC plots of the performance for (D) retrospective test sets; and (E) prospective test set (total cohort and individual hospital sites as indicated). (F) Overall performance metrics of prediction of the neural network model for five specific histopathological types per site. Graduated colors in the shaded bar indicate accuracy levels. Numbers in the boxes indicate the number of correctly identified samples/total sample number.

**Figure 4.**
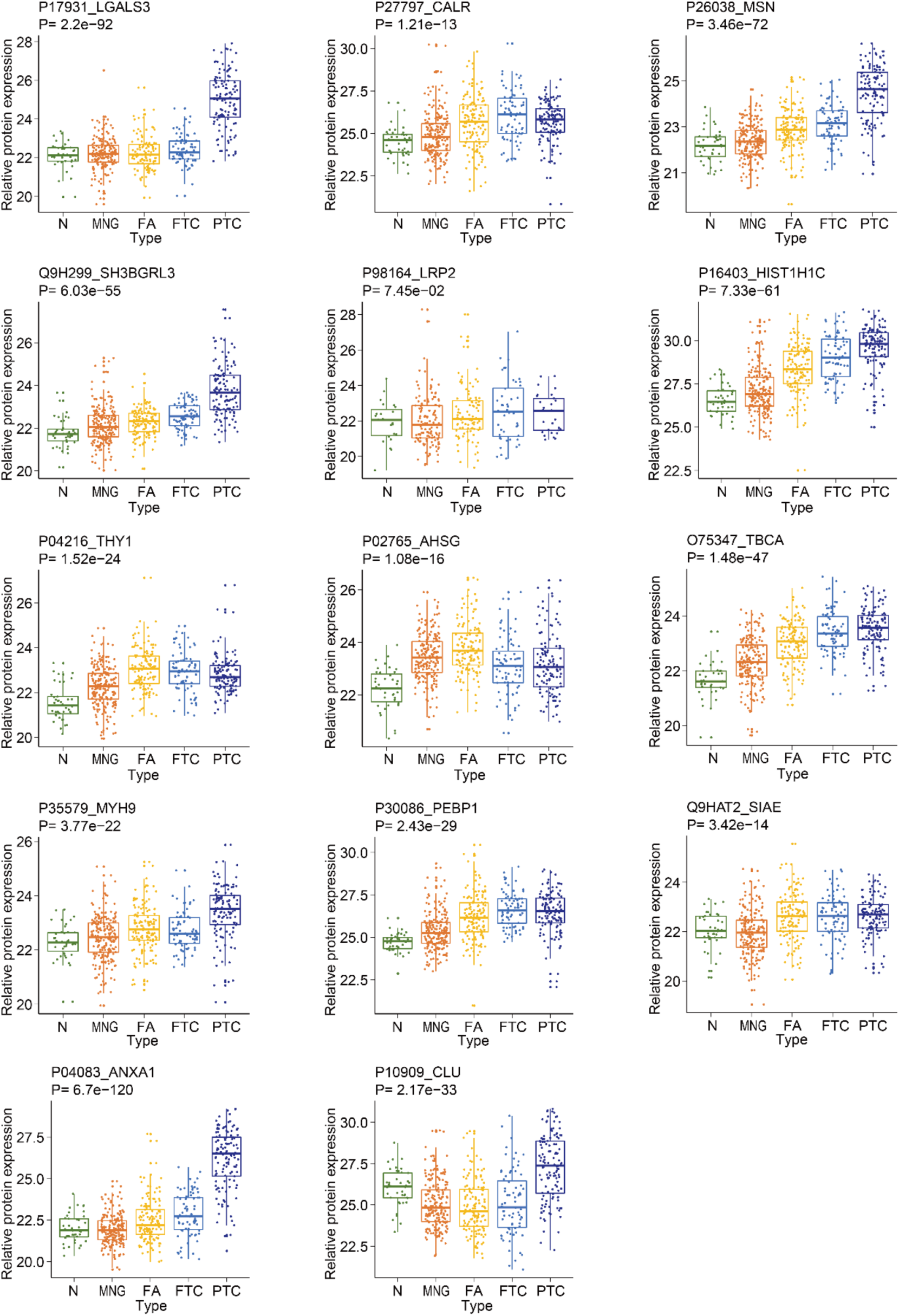
Protein expression plots for 14 selected protein features in the five types of thyroid tissues in the discovery cohort. Y-axis shows Log2 values of protein expression intensity and x axis indicates tissue type. P-value was calculated by one-way ANOVA. Further protein details are listed alongside each plot.

We next compared seven different machine learning models for classification using these 14 selected proteins. Receiver operating characteristics (ROC) plots showed the model based on artificial neural networks described here achieved the highest area under the curve (AUC) value of 0.95 and accuracy of 0.91 (Figure 3B). Using the 14 protein features in our established neural network model, each specimen was re-classified into benign or malignant in the 500 samples in the discovery cohort. The model comprised a ‘feature extraction sub-model’ which extracts and maps features from protein data into a feature vector in latent space, and a ‘classification sub-model’ which assigns a score (from 0 to 1) to the feature vector indicating the likelihood of malignancy for each sample. The ‘feature extraction sub-model’ was trained using contrastive loss function, while cross-entropy loss function was used to train the classification sub-model. Details of the neural network model are described in Supplementary Methods. We validated this model using the 79 internal validation samples (Table S4; Figure S3A-S3B). ROC plot showed that our model achieved an AUC value of 0.96 for the training dataset (n=500) used to derive this algorithm and 0.96 for the cross-validation dataset (n=79) (Figure 3C). t-SNE plot of the feature latent space showed remarkable separation between malignant and benign tissue using the 14-protein panel.

### Performance of the protein classifier

In order to validate this 14-protein model in an independent patient cohort, we first analyzed 288 pathologist-reviewed FFPE tissues (n = 271 patients) from three high-volume hospitals, comprising 144 benign and 144 malignant tissue samples. To ensure rigorous validation, the diagnoses were blinded during data acquisition and analyses. Each sample was analyzed using the PCT-DIA workflow in technical duplicates. Analysis of the resulting 576 DIA maps identified 3,527 high-confidence proteotypic proteins (Table S3). Individual ROC plots for samples from each of the three hospitals using the 14-protein model showed AUC of >0.91 for the retrospective FFPE samples (Figure 3D). Both scatter and t-SNE plots demonstrated distinct separation between benign and malignant thyroid tissues (Figure 3D; Table S4), although there were variations between individual sites (Figure S3C). The overall sensitivity and specificity were 90% and 91%, respectively, with negative-(NPV) and positive-predictive values (PPV) of 90% and 91%, respectively. Further details are provided in Table 3.

**Table 3.** Performance of the protein classifier.

| Center | Retrospective test sets |  |  | All retrospective study | Prospective test set | All |
| --- | --- | --- | --- | --- | --- | --- |
|  | ZY | ZE | DL |  | FNA |  |
| Sample type | FFPE | FFPE | FFPE | FFPE | Biopsy | FFPE+Biopsy |
| Malignant nodules No. | 35 | 61 | 48 | 144 | 46 | 190 |
| Benign nodules No. | 19 | 79 | 46 | 144 | 18 | 162 |
| Prevalence<sup>a</sup> | 64.81 (53.62 - 74.59) | 43.57 (36.84 - 50.54) | 51.06 (42.65 - 59.42) | 50.00 (45.16 - 54.84) | 71.88 (61.86 - 80.11) | 53.98 (49.58 - 58.31) |
| Predict M/M<sup>b</sup> | 31 | 58 | 41 | 130 | 40 | 170 |
| Predict B/B<sup>c</sup> | 17 | 71 | 43 | 131 | 16 | 147 |
| Sensitivity | 88.57 (76.80 - 94.78) | 95.08 (88.29 - 98.02) | 85.42 (75.12 - 91.91) | 90.28 (85.43 - 93.63) | 86.96 (76.67 - 93.12) | 89.47 (85.23 - 92.61) |
| Specificity | 89.47 (72.59 - 96.46) | 89.87 (82.88 - 94.21) | 93.48 (84.73 - 97.37) | 90.97 (86.23 - 94.19) | 88.89 (71.30 - 96.26) | 90.74 (86.28 - 93.85) |
| PPV | 93.94 (86.86 - 97.76) | 87.88 (82.57 - 91.70) | 93.18 (88.11 - 96.67) | 90.91 (87.79 - 93.39) | 95.24 (88.81 - 98.12) | 91.89 (89.01 - 93.87) |
| NPV | 80.95 (71.33 - 88.61) | 95.95 (91.91 - 97.77) | 86.00 (79.27 - 91.03) | 90.34 (87.01 - 92.79) | 72.73 (63.51 - 81.45) | 88.02 (84.92 - 90.63) |
| Accuracy | 88.89 (79.89 - 94.16) | 92.14 (87.54 - 95.14) | 89.36 (82.96 - 93.54) | 90.62 (87.40 - 93.09) | 87.50 (79.12 - 92.82) | 90.06 (87.11 - 92.39) |
| Youden's index | 0.78 | 0.85 | 0.74 | 0.81 | 0.76 | 0.80 |

Each value was calculated to 95% Wilson confidence intervals.
<sup>a</sup>The ratio of carcinoma in total nodules.
<sup>b</sup>The number of benign nodules identified as benign.
<sup>c</sup>The number of malignant nodules identified as malignant.

Given that the eventual objectives were to apply this analysis to pre-surgical FNA biopsies, we extended validation to a separate prospective cohort of 64 FNA samples (n=62 patients) obtained from a fourth independent clinical center. Even from these small amounts of FNA biopsy tissues, we were able to generate a high-quality protein matrix containing 3,310 proteotypic proteins using PCT-DIA technology (Table S3). The model achieved an AUC value of 0.89 (Figure 3E) and correctly identified 56 of 64 samples with sensitivity, specificity, PPV and NPV of 87%, 89%, 95% and 73%, respectively (Table 3). The predictive accuracy for all subtypes exceeded 91% except FA (89%) and FTC (83%) (Figure 3F), likely due to similarity of histology (and potential biological overlap) between these two pathologies.

### Biological insights on thyroid tumor subtypes

We next asked whether the proteomic data could be used to reveal biological insights of specific subtypes of thyroid neoplasms. In recent years, differentiated thyroid cancers have been further sub-classified based on specific morphological features or their expected clinical course. Hürthle cell adenomas and carcinomas are deemed as distinct entities, with the latter demonstrating a higher propensity for metastasis (Ganly et al., 2018; Gopal et al., 2018). The proteomic data of this study showed these to be well-resolved from other neoplasms, even from the closely related FA and FTC subtypes (Figure 5A; Figure S4A-S4C). Hürthle cell tumors are known for their oncocytic morphology and increased glucose uptake in FDG-PET scans (Grani et al., 2018), and indeed, our data showed that 91 of 109 proteins substantially elevated (fold change > 2 and adjusted p-value < 0.01) compared to follicular neoplasms are mitochondrial proteins participating in multiple metabolic pathways including the TCA cycle and oxidative phosphorylation (Figure 5B). Our data therefore uncovered biochemical processes contributing to the elevated metabolism of Hürthle cell tumors. Compared to the other four complexes in the oxidative phosphorylation pathway, the most strongly upregulated proteins (7/16) were in complex V which catalyzes ATP synthesis and potentially enhances tumor growth.

**Figure 5.**
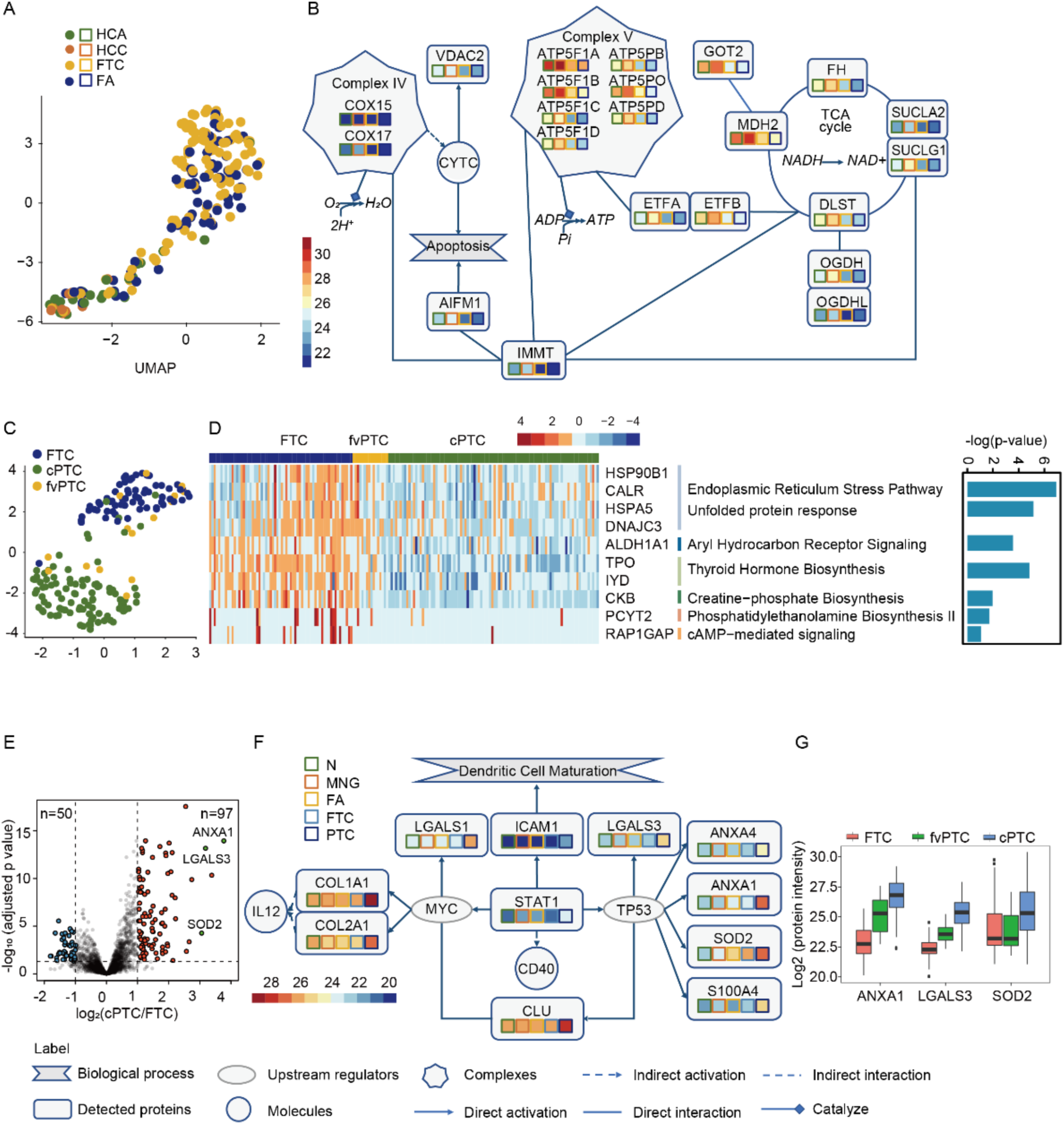
Biological insights of thyroid tumor subtypes based on proteotypic data. UMAP plot for 109 proteins distinguishing Hürthle cell from other follicular neoplasms. (B) Network map showing expression of key mitochondrial proteins implicated in Hürthle cell neoplasms. (C) UMAP plot for 175 proteins distinguishing FTC from cPTC, with fvPTC as an intermediate phenotype. (D) Heatmap showing differentially expressed proteins in FTC compared with fvPTC and cPTC, with pathways as indicated. X-axis of the vertical bar plot indicates −log2(p-value) based on right-tailed Fisher’s exact test from the IPA database. (E) Volcano plot showing 50 up-regulated proteins in FTC and 97 up-regulated proteins in cPTC with two-fold-change cutoff and adjusted p-value threshold less than 0.01. (F) Proteins in the cPTC network participate in immune pathways and oncogenesis (MYC and TP53 pathways). (G) Box plots showing three proteins specifically overexpressed in cPTC compared to other histological subtypes.

Follicular-variant papillary thyroid cancers (fvPTC) is a subtype with mixed morphology, and we therefore examined specific differences between FTC, classical papillary cancers (cPTC) and fvPTC. There were no significant proteotypic differences between FTC and fvPTC (Figure S4D). However, 45 proteins were differentially regulated in fvPTC compared to cPTC (Figure S4D; proteins listed in Table S5). Our proteotypic data showed that fvPTC overlapped with both FTC and cPTC, but resembled FTC more closely, indicating that fvPTC is potentially an intermediate entity between FTC and cPTC (Figure 5C). This is consistent with genomic classifiers which suggest that FTC and fvPTC share common alterations including in the RAS pathway (Agrawal et al., 2014). Compared to cPTC, 50 proteins upregulated in histologically distinct FTC participate in multiple biological processes including endoplasmic reticulum stress and the unfolded protein response (Figure 5D, S4E), both of which are implicated in bypassing Ras-driven oncogenic senescence (Blazanin et al., 2017). The signaling network involving key upregulated proteins in FTC is shown in Figure S4F. In contrast, the 97 proteins upregulated in cPTC compared to FTC (Figure 5E) mapped to major oncogenic (TP53, MYC), dendritic cell maturation pathways and inflammatory response, suggesting that canonical nocogenic pathways and inflammation are involved in the pathogenesis of cPTC, which has been associated with thyroid inflammation (Figure 5F). Consistent with this suggestion, three proteins involved in inflammation, ANXA1, LGALS3 and SOD2, showed greatest fold-change among proteins that mapped to the TP53- and MYC-related networks in cPTC (Figure 5G). Intercellular adhesion molecule 1 (ICAM-1) and signal transducer and activator of transcription 1 (STAT1) are over-expressed in cPTC and are potential immune-modulating targets.

## DISCUSSION

Molecular diagnostics for thyroid nodules have been limited to nucleic acid-based testing thus far due to the feasibility of analyzing small clinical samples and the increasing affordability of next-generation sequencing. Several nucleic acid-based tests are commercially available through central-lab testing, their performance in clinic is suboptimal in terms of specificity, especially in malignancies with low mutational burden as rigorously examined by Wang et al (Wang and Sosa, 2018). Since proteins are more stable than RNA in biopsy tissue samples (Shao et al., 2019), we posit that our protein panel can be developed as future point-of-care diagnostic tests through widely available techniques such as mass spectrometry and/or immunohistochemistry, as a complement to the nucleic acid-based test. The FFPE-PCT-DIA methodology used here was able to derive protein abundance data of 3,779 proteins in 931 samples, generating 2,421 DIA proteome data sets, including replicates. The pipeline generated the first repository of proteome data of various thyroid pathologies. This enabled artificial neural network analysis to mine large proteomic datasets for protein biomarkers of thyroid cancers. A panel of 14 proteins differentiated benign from malignant disease with diagnostic accuracy over 90% with sensitivity and specificity over 91% for retrospective test sets and accuracy of 89% for prospective FNA-derived test set. The fact that 12 out 14 of these have been previously implicated in thyroid physiology or pathology provides orthogonal validation for the inclusion of these proteins in our classifier. These metrics from proteomics data exhibited high degree of both sensitivity and specificity as shown in Table 3. Our method works for small tissue samples obtained from both FFPE tissues and FNA biopsies, making it broadly applicable for standard clinical practice, bypassing RNA assays due to the fragility of RNA integrity.

Despite the accuracy in distinguishing benign from malignant thyroid nodules, the major limitation for most algorithms is distinguishing FA from FTC. Indeed, the proteotype data presented here even suggests that this entity may represent a continuum of disease, in which differences exist at the extremes of phenotypes, but with significant overlap in-between. Alternatively, the overlapping benign categories may represent precursor lesions diagnosed prior to overt capsular or vascular invasion, even though pre-requisite conditions for invasion are already present in the tumors.

In conclusion, we present the first protein-based artificial neural networks classifier for classifying thyroid nodules. Although this test has been retrospectively validated in three clinical centers and prospectively validated in a fourth independent center, further validation using FNA biopsy specimens from larger prospective cohorts are required. This large-scale thyroid proteome profile of 931 thyroid nodules coupled with artificial neural networks demonstrates the power of a protein-based disease classifier with rapid potential to be translated into clinical practice. The thus established protein classifiers may complement nucleic acid-based tests in multiple clinical applications. Expanding this robust workflow to other carefully curated clinical cohorts may offer unprecedented opportunities to gain fundamental insights into molecular pathogenesis of diseases and address critical unmet clinical needs beyond thyroid cancer.

## Data Availability

All data are available in the manuscript or the supplementary material. MS raw data were deposited in iProX (IPX000444000). All the data will be publicly released upon publication.

## ACKNOWLEDGEMENTS

This work was supported by grants from National Natural Science Foundation of China (81972492) and National Natural Science Fund for Young Scholars (21904107), Zhejiang Provincial Natural Science Foundation for Distinguished Young Scholars (LR19C050001), Hangzhou Agriculture and Society Advancement Program (20190101A04). Further support for this project was obtained from the National Cancer Centre Research Fund (Peter Fu Program) and National Medical Research Council Clinician-Scientist Award (NMRC/CSAINV/011/2016), both in Singapore. The prospective study was supported by National Natural Science Foundation of China (81902726), China Postdoctoral Science Foundation (NO. 2018M641739), Natural Science Foundation of Liaoning Province (20180530090).

## AUTHOR CONTRIBUTIONS

T.G., N.G.I., O.L.K., S.Z.L., Y.Zhu and Y.S. designed the project. SS performed central pathology review of the discovery set. H. Z. collected the samples for prospective test set. L.C., X.T., Y.Zhao, G.W., J.X., Z.W., W.S., Y.Q., Y.H., L.W. and S.Z. procured and annotated the Chinese thyroid tissue samples, while SS, SM, TKHL, SY, SMFSA, SSL, BH procured and annotated the Singapore tissue samples. Y.S., W.L., X.C., X.Z., Q.X., H.G., X.Z. and K. X. performed the experiments. Y.S., W.L., W.W., H.C., T.Z. and Q.Z. conducted proteomic data analysis. Z.Z. and S.Z.L. performed the learning analysis. Y.S., N.G.I., O.L.K. and T.G. wrote the manuscript with inputs from all co-authors. T.G., N.G.I., O.L.K. and S.Z.L. supervised the project.

## MATERIALS AND METHODS

### Patients and tissue samples

Tissue cores (1 mm diameter, approximate weight 0.6 mg, including was) were punched from blocks of formalin-fixed paraffin embedded (FFPE) thyroid tissues obtained from four clinical centers in Singapore and China spanning 2011-2019, with ethics approval of each hospital.

The discovery sample set of 579 thyroid nodules from 578 patients comprised follicular adenomas (FA, 137 cases), multinodular goiters (MNG, 203 cases), papillary thyroid carcinoma (PTC, 124 cases) and follicular thyroid carcinoma (FTC, 75 cases) from the Singapore General Hospital. Hematoxylin and eosin-stained slides from tissue blocks of each patient were reviewed by an experienced histopathologist who marked out the disease region for tissue coring. Normal thyroid tissues (N, 40 cases) were taken from cases of laryngectomy or pharyngo-laryngo-esophagectomy in which the thyroid gland was surgically removed incidental to radical surgery. These patients had no history of thyroid disease, prior chemotherapy or radiation. Three tissue cores were made for each case as biological replicates.

For multi-center blinded test sets, we firstly analyzed a total of 288 FFPE tissue cores from 271 cases composed of 44 MNG, 84 FA, 52 FTC and 92 PTC cases in retrospective test sets. Sixteen cores were of adjacent normal thyroid tissue (N). A single punch was made from each case. Furthermore, we tested 64 fine needle aspiration biopsies containing 13 MNG, 5 FA and 46 PTC cases in prospective test set. Histological diagnoses of these cores were blinded during the entire validation workflow of sample processing, mass spectrometry analysis and predictive data analysis.

Cases of microcarcinoma, extensive thyroiditis and/or inflammation were excluded from the discovery sample set.

### Batch design

To minimize batch effects among different lots of analyzed samples, 1,725 FFPE cores from 579 thyroid nodules and 56 peptides extracted from cores selected as technical replicates from the discovery sample set were randomly distributed into 121 batches. Each batch contained 15 thyroid samples, one mouse liver sample as quality control (QC) for PCT and one thyroid pooled sample containing all five types of thyroid tissues for mass spectrometry (MS). In this discovery phase analysis, the histopathology diagnosis of each tissue core was known from which models of data segregation were established.

In the test phase analysis, 288 FFPE cores were analyzed in technical duplicates for a total of 576 samples in 44 batches for retrospective test sets and 64 fine needle biopsies in 5 batches for prospective test set (Figure S1).

### Dewaxing, rehydration and hydrolysis of FFPE tissues

For each case in the discovery sample set, three biological replicates of FFPE tissue cores were processed. Sample weights were recorded before dewaxing in heptane (Sigma) and successively in 100% ethanol (Sigma), 90% ethanol, 75% ethanol at room temperature. Formic acid (0.1%) (Sigma) was added next for achieve C-O hydrolysis of protein methylol products and then washed with 100 mM Tris-HCl (pH10, Sigma) to establish conditions for base hydrolysis at 95°C. The sample was then snap cooled to 4°C.

### Tissue lysis, protein extraction and protein digestion

Dewaxed samples were lysed in 6M urea (Sigma) and 2M thiourea (Sigma) using pressure cycling technology (PCT) programmed for 90 cycles of 25 s at 45,000 p.s.i. and 10 s at ambient pressure at 30°C. After lysis, 10 mM Tris(2-carboxyethyl) phosphine hydrochloride and 40 mM iodoacetamide were simultaneously added to the solution and incubated in the dark with gentle vortexing for 30 min, after which lysC (Hualishi Scientific) was added at a ratio of 40:1 (protein to lysC). PCT-assisted lysC digestion was performed with the following setting: 45 cycles of 50 s at 20,000 p.s.i. and 10 s at ambient pressure at 30°C. Final tryptic digestion was performed at a ratio of 40:1 (protein to trypsin) by PCT with the following setting: 90 cycles of 50 s at 20,000 p.s.i. and 10 s at ambient pressure at 30°C. Peptides were desalted before LC-MS analysis.

### DIA library construction

DIA (data-independent acquisition) library was built as described previously (Guo et al., 2015; Schubert et al., 2015). To build the spectral library for analyzing DIA files from thyroid tissue samples, we collected tissue samples from the all five types of patient groups into a single pool. The tissue samples were either fractionated into six fractions using strong cation exchange (SCX) or processed with PCT-assisted lysis and in-solution digestion, or PCT-assisted lysis and PCT-assisted digestion. Finally, peptides were desalted on C18 columns.

Desalted peptides were separated by Ultimate 3000 nanoLC-MS/MS system (Dionex LC-Packing, Amsterdam, The Netherlands) equipped with 15 cm* 75 μm ID fused silica column custom packed with 1.9 μm 100 Å C18 AQUA. Peptides were separated on a 120 min (148 min inject-to-inject) LC gradient at 300 nL/min in a 3-25% linear gradient (buffer A: 2% acetonitrile,0.1% formic acid; buffer B: 98% acetonitrile, 0.1% formic acid). Eluted peptides were ionized at a potential of +2.0 kV into Q Exactive HF mass spectrometer (Thermo Fisher, Bremen, Germany). The full MS was measured at resolution 60,000 (at *m/z* 200) in an Orbitrap using an AGC target value of 3E6 charges. The top 20 peptide signals (charge-states +2 and higher) were submitted to fragmentation in HCD cell (higher-energy collision, 27% normalized collision energy), and then transferred to Orbitrap for MS/MS analysis. MS/MS spectra were acquired at resolution 30,000 (at *m/z* 200) in Orbitrap using an AGC target value of 1E5 charges, a maxIT of 80 ms and the dynamic exclusion time was 30 s.

In total, we acquired 52 DDA files including 20 SCX fraction files, 18 in-solution digestion files and 14 PCT-assisted digestion files on a QE-HF mass spectrometer in DDA mode (Table S2). All DDA files were centroided and converted to mzXML using msConvert from ProteoWizard with parameters ‘—mzXML --filter “peakPicking true [1,2]”‘. We analyzed DDA files using pFind version 3.1.3 with SwissProt fasta files (20,269 protein sequences), and identified 47,229 transition groups, 33,802 peptides, 5,190 protein groups and 4,048 proteotypic proteins.

### DIA-MS data analysis

Peptides were separated by using Ultimate 3000 or nanoLC-MS/MS system (DIONEX UltiMate 3000 RSLCnano System, Thermo Fisher Scientific™, San Jose, USA) equipped with 15cm*75 μm ID fused silica column custom packed with 1.9 μm 120 Å C18 aqua. Peptides were separated on a 45 min (68 min inject-to-inject) LC gradient at 300 nL/min in a 3-25% linear gradient (buffer A: 2% acetonitrile,0.1% formic acid; buffer B: 98% acetonitrile, 0.1% formic acid). Peptides eluted from analytical columns were ionized at a potential +2.0 kV into Q Exactive HF mass spectrometer (Q Exactive Hybrid Quadrupole-Orbitrap, Thermo Fisher Scientific™, San Jose, USA). A full MS scan was acquired analyzing 390-1010 *m/z* at resolution 60,000 (at *m/z* 200) in the Orbitrap using an AGC target value of 3E6 charges and maximum injection time of 100 ms. After the full MS scan, 24 MS/MS scans were acquired, each with a 30,000 resolution (at m/z 200), AGC target value of 1E6 charges, normalized collision energy of 27%, with the default charge state set to 2, maximum injection time set to auto. The cycle of 24 MS/MS scans (center of isolation window) with three kinds of wide isolation window was as follows (m/z): 410, 430, 450, 470, 490, 510, 530, 550, 570, 590, 610, 630, 650, 670, 690, 710, 730, 770, 790, 820, 860, 910, 970. The entire cycle of MS and MS/MS scan acquisition took approximately 3 seconds and was repeated throughout the LC/MS analysis. DIA raw files were analyzed using OpenSWATH v2.0 (Rost et al., 2014).

### Data quality control

We first assessed data quality by analyzing samples. The QC samples were mouse liver samples (PCT-QC) and pooled thyroid samples (DIA-QC) in each batch. Additional QC samples were analyzed as technical replicates for MS. Biological replicates were also analyzed to determine the extent of heterogeneity of thyroid diseases. Reproducibility of spiked-in mouse liver samples and thyroid pooled samples showed that PCT and MS instruments were stable during data acquisition (Figure S2A and B), with median coefficient of variance (CV) less than 0.03. MS data of 56 randomly selected paired thyroid samples in the discovery cohort had a median Spearman correlation coefficient of 0.9 (Figure S2C). CV for proteins in biological replicates was 0.2, indicating minimal tissue heterogeneity in the biology of thyroid disease (Figure S2D). Spearman correlations of four pooled samples and mouse liver samples are higher than 0.9 in prospective test set (Figure S2E and F)

### Development of neural network classifier

A neural network classifier is developed to classify any given proteome data sample (of 14 features) into one of two classes, benign (B) or malignant (M), so as to achieve the best accuracy. This comprises 4 stages:

1. Protein data preprocessing;
2. Protein feature selection using a genetic algorithm (GA);
3. Learning a neural network (DNN)-based classifier;
4. Using the trained DNN for sample classification.

The following details the algorithms for the 4 stages.

Stage 1: Data preprocessing

Four datasets from 5 cohorts, denoted SG, ZY, ZE, DL, FNA, were used for the development of the DNN model. There, 500 random samples from the SG cohort were used to train the model, 79 from the same cohort were used for internal validation, and data from the ZY, ZE, DL and FNA cohorts were used for external validation. Note that the external validation datasets are different those of training and internal validation. A data sample in these datasets consists of 3,708 proteins.

The preprocessing consists of two steps: (1) missing value imputation and (2) normalization. Missing values are inevitably a feature of protein intensity data. Considering most missing values occur when the protein content is below a detection threshold, the imputation is done by filling in all the missing values with 0.8\**D*_*min*_, where *D*_*min*_ is the minimum of all feature values in the training and validation sets and for this work, *D*_*min*_ = 13.

Thereafter, for each feature *D* after the imputation step, the mean *μ* and *σ* variance of that feature are estimated from the training and validation sets, and the feature of every training sample is normalized according to the following

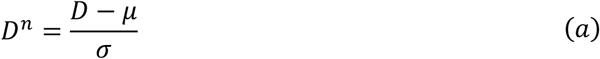

where *D*^*n*^ is the normalized feature. The obtained *μ* and *σ* are applied to the corresponding protein features in the testing phase afterwards.

Python’s pandas library is used to complete data preprocessing. Stage 2: Features selection

Feature selection is needed for two reasons: (1) most of the 3,708 proteins are useless or may be conflicting for the classification and they should be removed from consideration in DNN-based classifier learning; (2) it is desired to minimize the number of proteins in clinical applications. It is done in two steps (Figure S5A).

The first step is feature screening. Out of the initial 3,708 protein features, 521 were selected from differentially expressed protein of benign and malignant samples in the SG dataset and from published literatures related to thyroid or thyroid cancer. Among the 521, 64 did not appear in our datasets and are excluded, with 477 candidates remaining. Further, if such a protein has a deletion rate greater than 45%, then it is removed, with 275 proteins left. Even further, a pair of protein are removed if the absolute value of the Pearson correlation between them is less than 0.1, yielding the 243 candidate proteins.

As the second step, a combinatorial optimization is performed to select a best combination of 14 proteins from the 243. While no algorithms can guarantee a globally optimal solution with efficiency, here a genetic algorithm (GA)(Mitchell, 1998) was used to find the best 14 proteins.

In GA, evolutionary operation (crossover, mutation, and selection operations) are used to generate new protein feature combinations from existing protein features combinations. At every iteration, the GA eliminates the low fitness combinations, and generates new combinations based on the remaining high fitness combinations.

A fitness value is calculated for each candidate combination solution (gene chain). For gene chain *C*, it is defined as

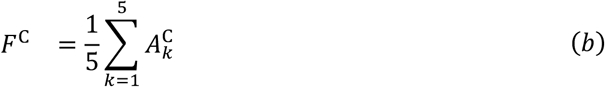

where 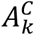 is the accuracy of the *k*-th cross-validation(Kohavi, 1995), which is computed from the difference between the output of the classifier and the true label.

Python’s deap library is used to complete features selection. Stage 3: Model training

The DNN classifier is a nonlinear function which takes a vector of 14 selected protein features as the input and produces a class label of either 1 (for malignant) or 0 (for benign) as the output. This consists of the following 3 steps: (1) model structure design; (2) loss function design; and (3) model training.

A multi-layer perceptron (MLP) structure is chosen for the DNN, shown in Figure S5B, The MLP model is divided into feature extraction sub-model and classification sub-model, trained in sequence as will be described below. The feature extraction sub-model extracts effective feature vectors (V_p_), and the classification sub-model completes classification diagnosis based on the classification information.

Once the DNN structure is defined, the parameterized nonlinear function is trained to achieve the best objective value as measured by a loss function. The loss function of the MLP is defined as

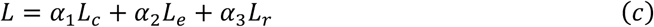

where *L*_*C*_ is a contrastive loss (Becht et al., 2018) for training the feature extraction sub-model, *L*_*e*_ is a cross-entropy loss for training the classification sub-model, *L*_*r*_ is an L2 regularization loss for reducing overfitting, and α_1_, α_2_, α_3_ > 0 are the weights.

The contrastive loss *L*_*C*_ is defined under the Siamese Network (SN) structure. The contrastive loss *L*_*C*_ is calculated as

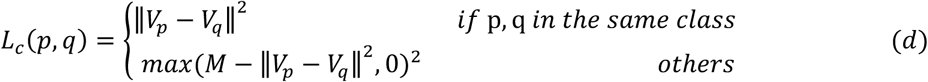

where feature vectors V_P_, V_*q*_ extracted from protein samples *X*_*p*_ and *X*_*q*_, ‖ V_p_− V_*q*_ ‖ is the distance, and *M* is the margin (M=1 in this work),. The contrastive loss encourages reduction of within-class feature scatter and increase of between-class feature scatter. The cross-entropy loss *L*_*e*_ is calculated as

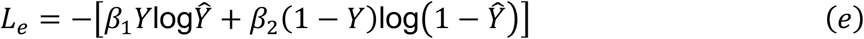

where *Y* is the real label of the patient, 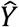 is the classification score predicted by classification sub-model, and β_1_, β_2_ are penalty parameters (β_1_ = 0.725, β_2_ = 1.275 in this work). The L2 regularizer is defined as the 2-norm of MLP weight a as follows

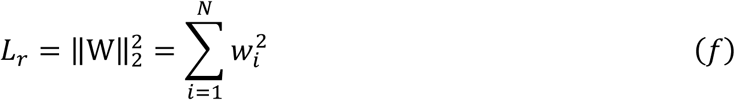

where *e* is the number of the layer.

The MLP training is done with the help of the training dataset (SG), in which the class label is provided for each sample as the supervisor to guide the training process. The MLP training process consists of two parts. The first part trains the feature extraction sub-model for 100 epochs, with α_1_ = 1, α_2_ = 0, α_3_ = 0.04 an *D* learning rate 1 × 10^−5^. The second part uses the cross-entropy loss training the classification sub-model for 900 epochs, with α_1_ = 0, α_2_ = 1, α_3_ = 0.029 an*D* learning rate 5 × 10^−4^, while the feature extraction sub-model parameters are not frozen for faster convergence. The obtained accuracy for the training set is 93%, and that for the validation set is 91%.

Python’s pytorch library is used to complete model training. Stage 4: Classification

The trained MLP is used as the classifier for diagnosis of unknown samples. Given the 14 features, the MLP outputs a classification score X*Y* between 0 and 1 and the class prediction P is calculated as follows

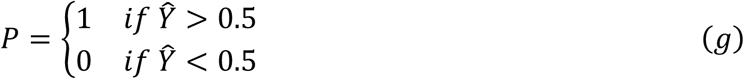

where P=0 means the tissue is predicted to be benign, and P=1 malignant.

### Statistical analysis

Statistical analysis was performed using R software (version 3.5.1) with pheatmap, UMAP and R package plot functions. CV was calculated as the ratio of the standard deviation to the mean. The prevalence for each cohort was based on the ratio of malignant to total tissues. Sensitivity, specificity, PPV and NPV values were calculated following the established methodology and each value was calculated with 95% Wilson confidence intervals (Steward et al., 2019). Biological insights were analyzed by Ingenuity Pathway Analysis (IPA version 49309495). P values were calculated by one-way ANOVA in the expression of 14 protein features.

---

**Figure S1.**
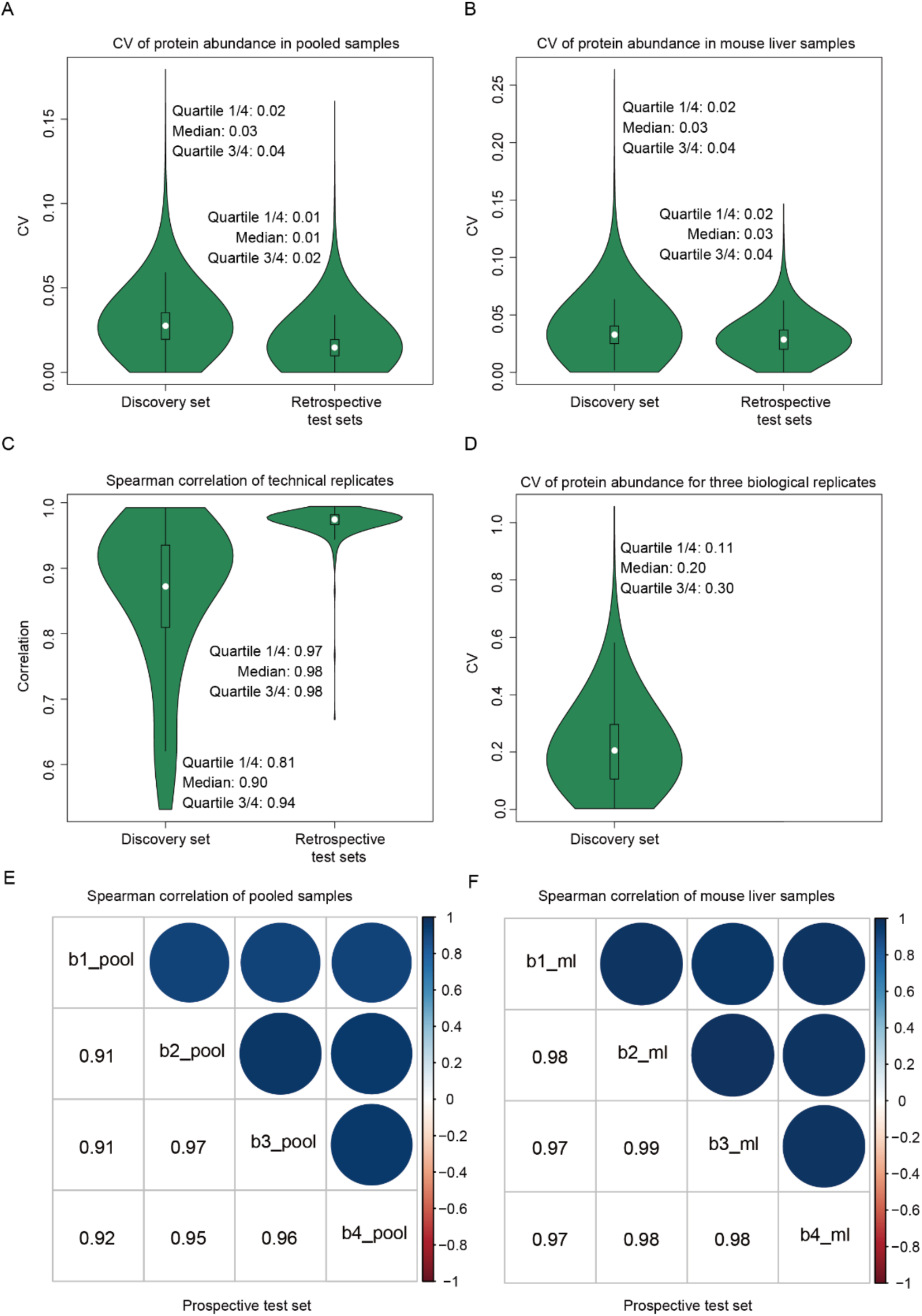
Data quality evaluation. (A) CV of identified protein numbers for 116 and 36 pooled thyroid samples in the discovery set and retrospective test sets, respectively. CV of identified protein numbers for 112 and 18 mouse liver samples in discovery and retrospective test sets, respectively. (C) Spearman correlation of paired technical replicates from 56 randomly selected thyroid samples in the discovery group and 288 in the retrospective test sets. (D) CV for the number of proteins in biological replicates of the discovery set. (E) Spearman correlation of four pooled samples in prospective test set. (F) Spearman correlation of four mouse liver samples in prospective test set.

**Figure S2.**
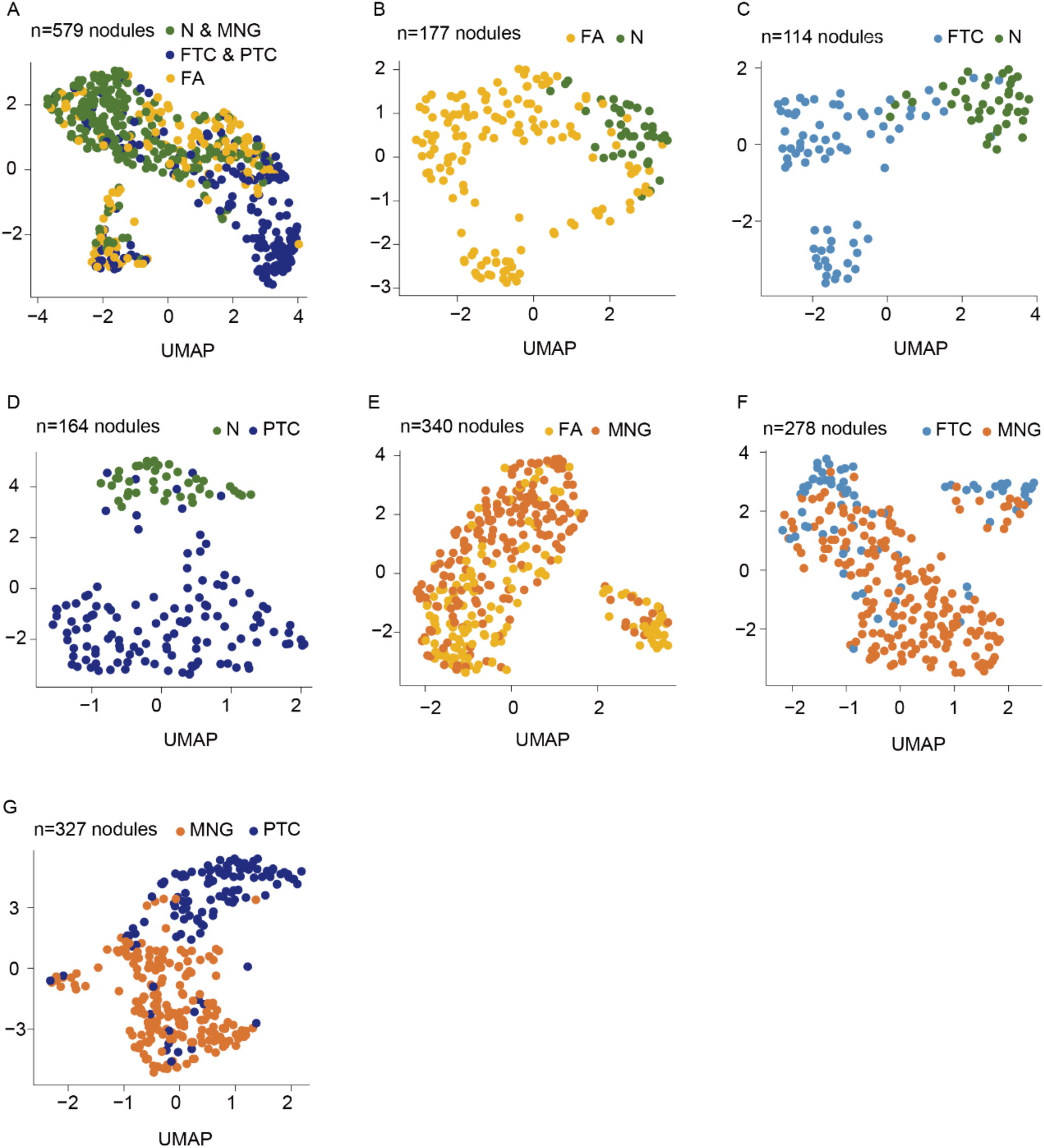
Uniform manifold approximation and projection (UMAP) analysis of five subtypes of thyroid tissues. 2,617 proteins for which missing value was less than 90% were used in data analysis. (A) All tissue types, showing FA distributed across benign (N and MNG) and malignant (FTC and PTC) tissues, (B) FA, (C) FTC, (D) PTC vs N, respectively, and (E) FA, (F) FTC and (G) PTC vs MNG, respectively. Normal tissue is generally well separated from all other lesional tissue, while MNG shows some overlap with FA, FTC and PTC.

**Figure S3.**
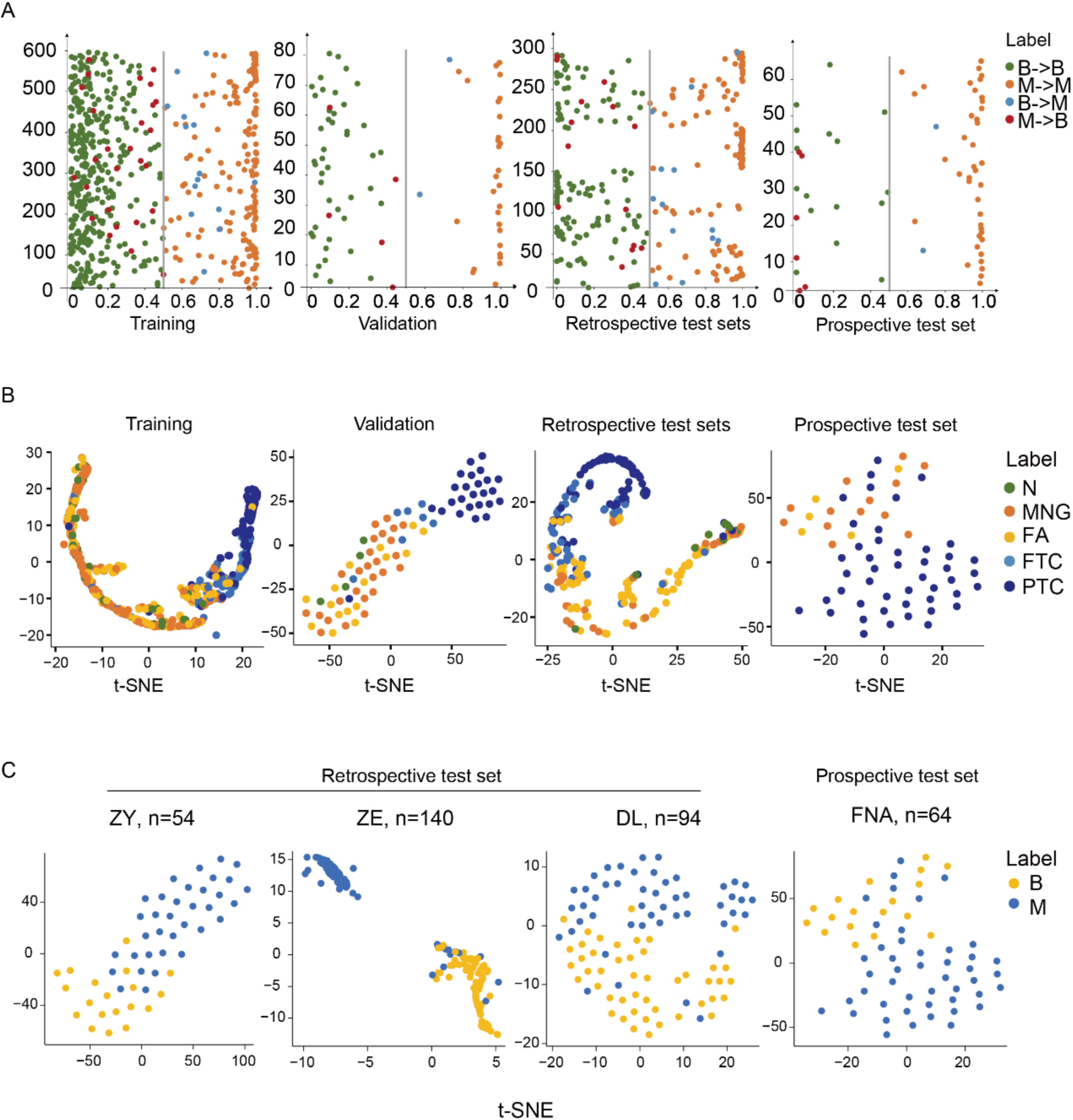
Cross-validation of the classifier on discovery set and performance on test sets. (A) Scatter diagram showing the predicted malignancy scores for discovery (training and validation), and tests sets (retrospective and prospective test sets), and the probability for each sample to be malignant. X axis indicates the probability of malignancy. Y axis represents the number of thyroid tissues in different sets. (B) t-SNE plots showing specific tissue types (benign and malignant) based on the 14 protein features in the training set, validation set, retrospective and prospective test sets, labeled by each of the 5 subtypes. (C) t-SNE plots based on the 14 proteins for each of the four clinical sites for the different test sets.

**Figure S4.**
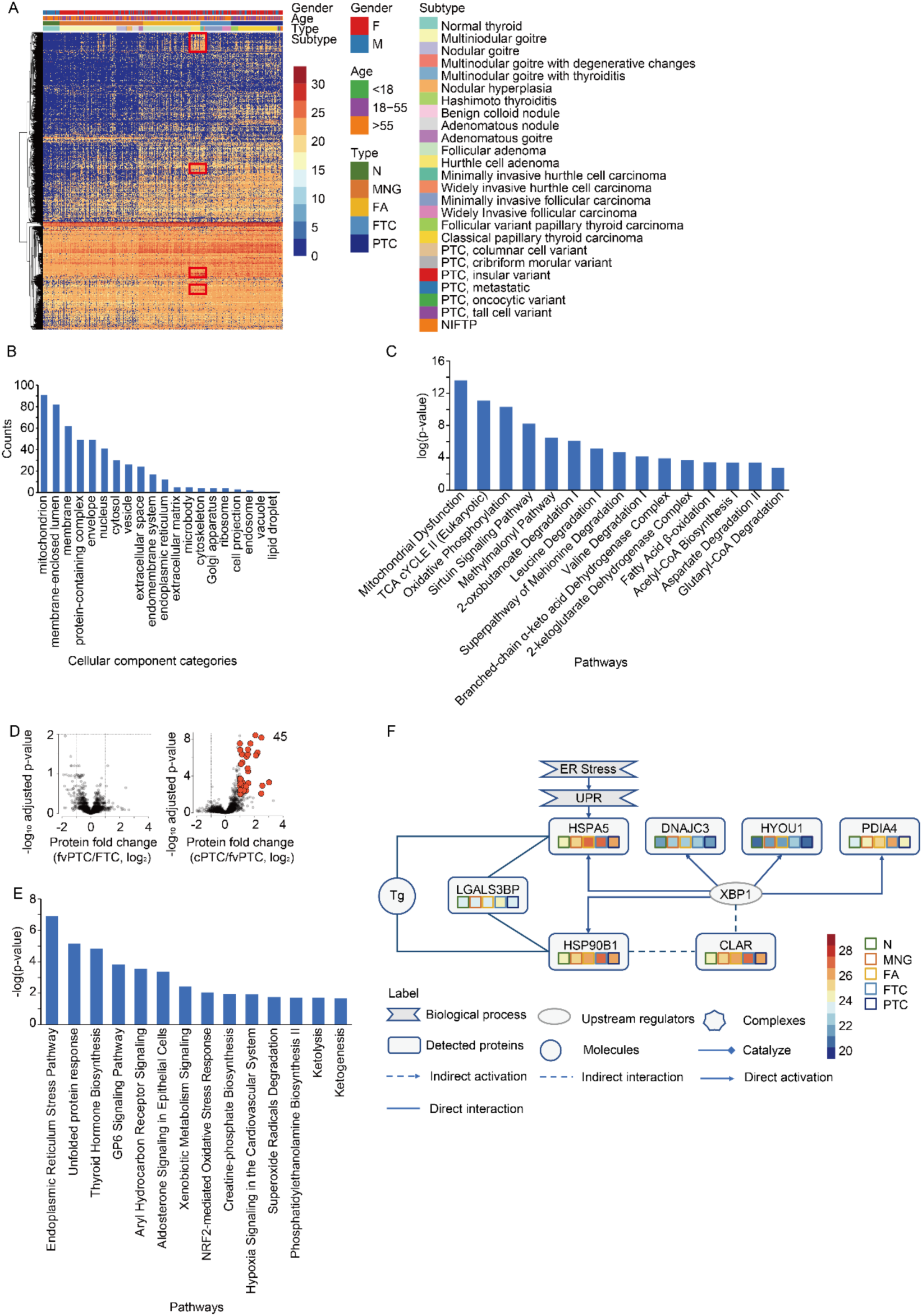
Biological insights into Hürthle cell tumors, follicular (FTC), classical papillary (cPTC) and follicular-variant PTC (fvPTC). (A) Heatmap showing proteotype expression of thyroid tissue samples highlighting differentially expressed proteins in the Hürthle cell neoplasm marked in red frames. The subtype label of heatmap were based on reviewed slides by experienced pathologist. (B) Graph showing cellular component analysis of the 109 over-expressed proteins in Hürthle cell neoplasms. Mitochondrial proteins are the most dominant group. (C) Graph showing enriched pathways based on 109 over-expressed proteins of Hürthle cell tumors by IPA analysis. Y-axis shows −log2(p-value) based on right-tailed Fisher’s exact test (IPA) based on the IPA database. (D) Volcano plots showing differentially expressed proteins between fvPTC vs FTC and cPTC vs fvPTC. Protein intensities used were from the average intensity of three biological replicates. We compared pair-wise groups, and highlighted proteins that were significantly different with two-fold-change cutoff and adjusted p-value threshold less than 0.01. (E) Pathway enrichment of 50 upregulated proteins in FTC compared with cPTC as analyzed by IPA. Y axis indicates −log2(p-value) based on right-tailed Fisher’s exact test based on the IPA database. (F) Network map showing overexpressed proteins in FTC.

**Figure S5.**
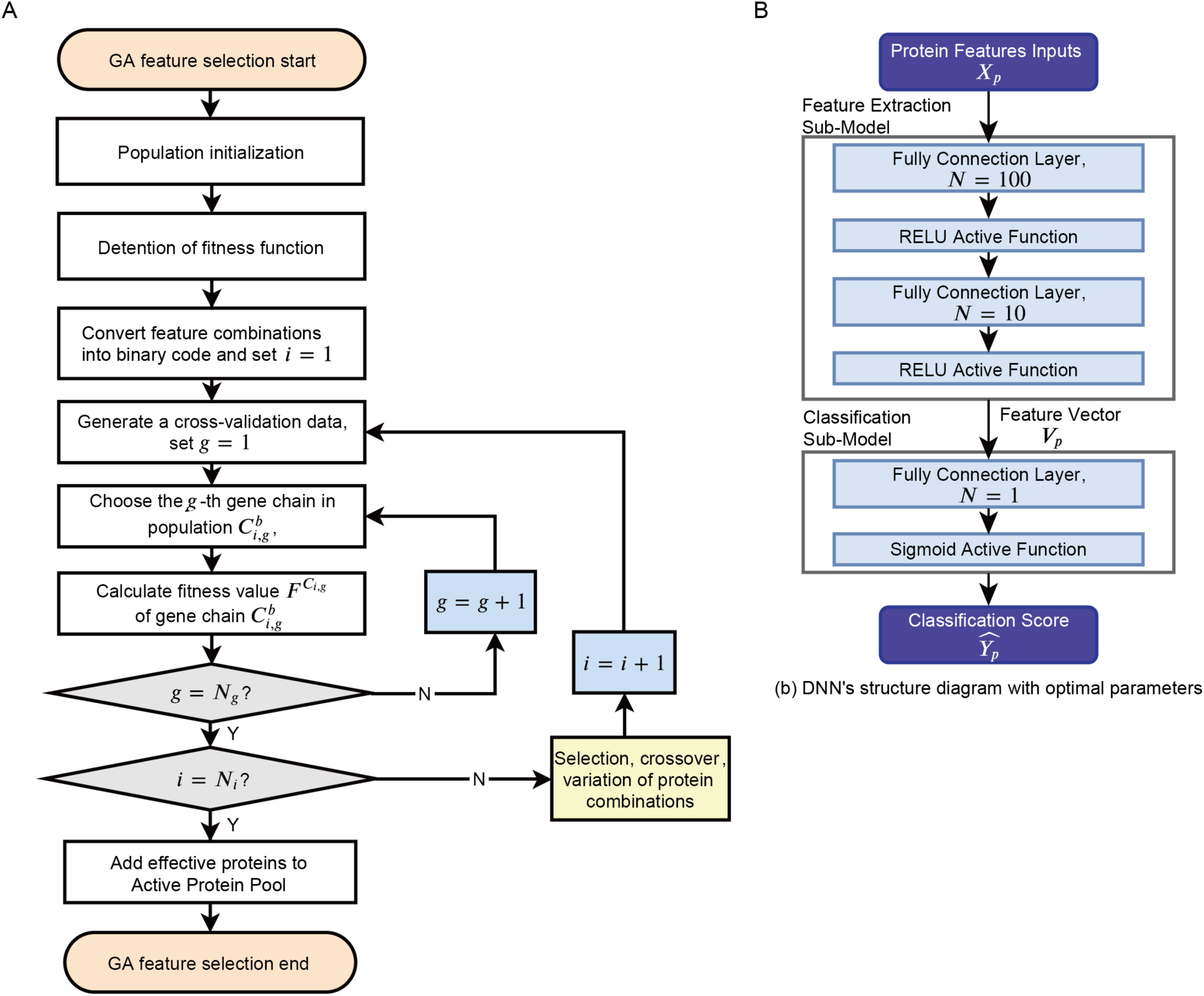
Structures of models. (A) Flow diagram of genetic algorithm for protein features selection, i is the index of iteration, g is the index of gene chains. N_g_ is the population size (number of the gene chains), N_i_ is the maximum number of iterations. 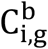 is the binary codes of i-th iteration g-th gene chian. 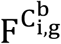 is the fitness function of 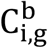. (B) DNN’s structure diagram with optimal parameters. p is the index of the patients, *X*_*p*_ is the protein feature inputting to the feature extraction sub-model, *V*_*p*_ is the output of the extraction sub-model, 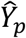 is the output of the classification sub-model and the classification score predict by DNN, *e* is the number of neurons in the layer.

